# Characterisation of disease progression in hantavirus haemorrhagic fever with renal syndrome

**DOI:** 10.64898/2026.06.15.26355676

**Authors:** Stuart Armstrong, Katarina Resman Rus, Nataša Knap, Efthyvoulos Drousiotis, Claire E. Eyers, Iain Buchan, Tatjana Avšič Županc, Julian A. Hiscox, Miša Korva

## Abstract

Hantaviruses can cause haemorrhagic fever with renal syndrome (HFRS). This is a clinically variable disease in which severe outcomes are hypothesized to arise from dysregulated host responses. To characterise this, longitudinal, label-free plasma proteomics was used to compare disease progression in a unique well-defined cohort of patients infected with either Dobrava virus (DOBV) or Puumala virus (PUUV) hantaviruses. Patients were stratified by clinical severity. The average viral load in the first available sample from hospitalized patients was higher in those who went on to have severe infection, and higher in patients infected with DOBV. There was marked separation of infected patients from controls across early, mid and late disease, including after viral RNA clearance, suggesting a sustained systemic host-response signature. Proteomic signatures were consistent with a strong acute-phase response in both mild and severe disease. There was evidence of activation of the adaptive humoral response at later stages. Hierarchical clustering identified severity-associated pathways linked to endothelial dysfunction, thrombocytopenia, vascular leakage and renal injury. These findings define a durable plasma proteomic signature of hantavirus disease and support a model in which severe HFRS is driven by persistent inflammatory, complement and platelet/coagulation pathway activation rather than viral burden alone.

## Introduction

Orthohantaviruses are enveloped, tri-segmented, negative-sense RNA viruses maintained in nature through persistent infection of small-mammal reservoirs, most commonly rodents. Human infection is typically incidental and occurs after inhalation of aerosolized excreta from infected reservoir hosts. Outbreaks are generally sporadic and related with the density of the rodent population. Pathogenic orthohantaviruses are classically associated with two overlapping clinical syndromes: haemorrhagic fever with renal syndrome (HFRS), reported mainly in Europe and Asia ^1, 2^ (Old World orthohantaviruses), and hantavirus cardiopulmonary syndrome, also termed hantavirus pulmonary syndrome (HCPS/HPS), reported mainly in the Americas ^3, 4^ (New World orthohantaviruses). HFRS is characterised by fever, thrombocytopenia, vascular leakage, hypotension, and acute kidney injury (Figure 1), whereas HCPS/HPS is dominated by rapidly progressive pulmonary oedema, respiratory failure, and cardiogenic shock ^3, 4, 5^. The disease severity varies substantially according to viral species and host factors. For example, Puumala virus (PUUV; *Orthohantavirus puumalaense*) infection is often associated with a milder form of HFRS, whereas Dobrava virus (DOBV; *Orthohantavirus dobravaense*) has been associated with severe disease ^6^. The reported case fatality for DOBV infection is around 10% ^7, 8^, whereas with PUUV infection this can be less than 1% ^9^.

**Figure 1.**
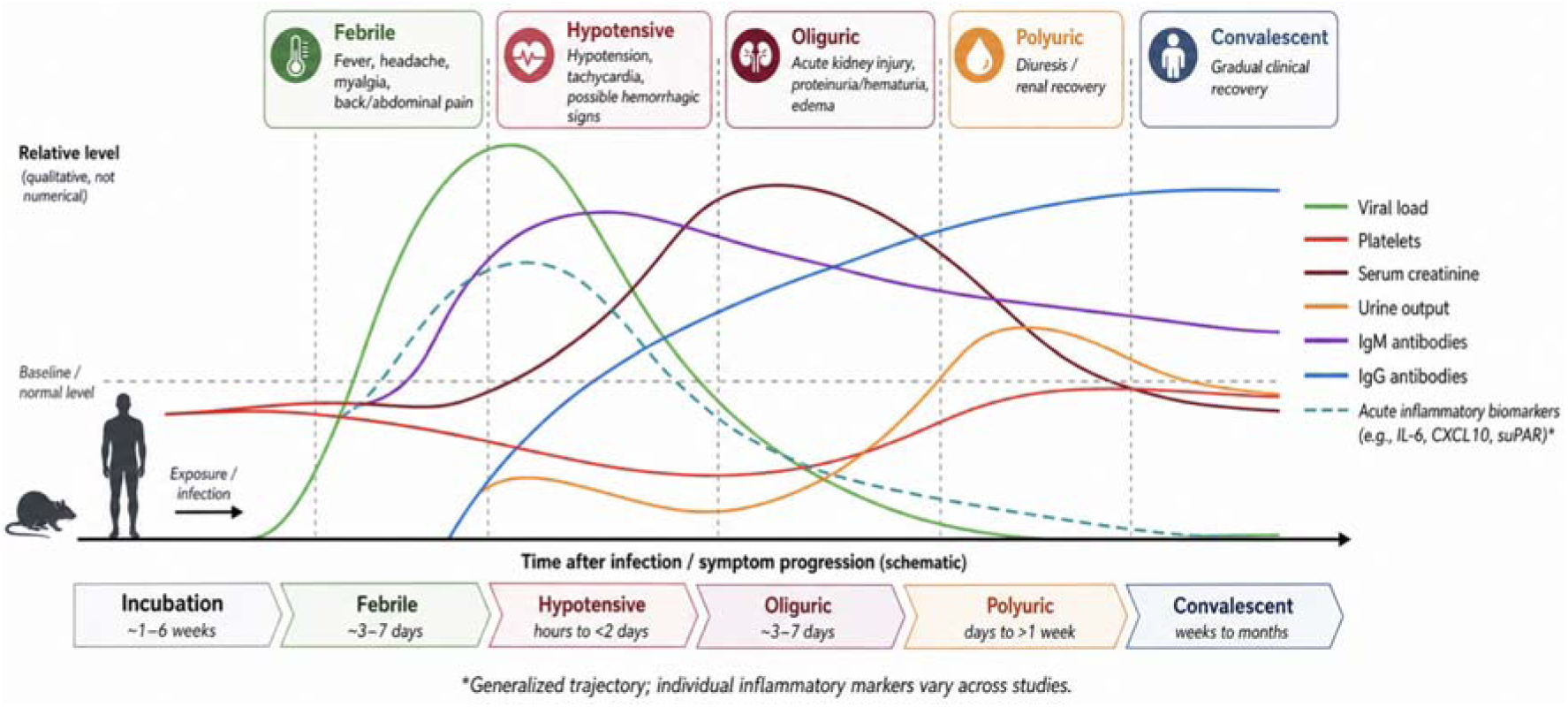
Schematic course of clinical phases and relative biomarker changes in patients with HFRS (not to scale). Exposure to infected material results in a febrile illness followed by potentially severe disease. Whilst viral load and inflammatory biomarkers may decrease, clinical symptoms may persist. Figure adapted from an earlier version we presented ^3^.

A central feature of hantavirus pathogenesis is vascular dysfunction in the relative absence of overt virus-induced cytopathic injury. Endothelial cells are major targets of infection, and pathogenic hantaviruses can infect the microvascular endothelium without directly lysing infected cells ^10, 11^. Instead, clinical disease is thought to reflect a multifactorial process involving endothelial activation, altered barrier function, platelet dysfunction, coagulation and complement abnormalities, and immune-mediated inflammation ^12^. Experimental studies have shown that hantavirus infection can sensitize endothelial cells to vascular permeability mediators such as vascular endothelial growth factor (VEGF), while other endothelial-stabilizing pathways may counteract this effect ^13^. These findings support the concept that hantavirus disease severity is driven not only by viral replication but also by the host response to infection.

The immune response to orthohantavirus infection is therefore central to both viral control and disease pathogenesis. Innate immune recognition induces type I interferon responses and pro-inflammatory cytokines and chemokines, while adaptive responses include virus-specific T-cell and antibody responses ^12, 14^. Several studies have suggested that strong cellular immune activation may contribute to endothelial dysfunction and tissue injury, particularly through recruitment and activation of cytotoxic T cells, natural killer cells, monocytes, and other inflammatory effector cells ^15^. Conversely, humoral immunity appears important for recovery, and neutralizing antibody responses have been associated with protection and improved outcomes in HCPS/HPS ^15, 16^. Thus, severe disease may reflect an imbalance in which protective antiviral immunity is accompanied by excessive or dysregulated inflammatory and endothelial responses.

Consistent with this model, circulating biomarkers of inflammation, endothelial activation, epithelial or tissue injury, coagulation, and organ dysfunction have been investigated as correlates of hantavirus severity and outcome. In HCPS/HPS, multiplex serum analyses have demonstrated broad systemic inflammatory activation, with several cytokines, chemokines, and tissue injury markers differing between fatal and non-fatal disease ^17, 18^. Interleukin-6 (IL-6) has been associated with disease severity, while intestinal fatty acid–binding protein, a marker of intestinal epithelial injury, was associated with fatal outcome ^17^. In PUUV infection, a range of immunoinflammatory biomarkers have been investigated in relation to HFRS severity, although many remain experimental and are not routinely used in clinical practice ^19^. Patients with HFRS exhibited elevated serum levels of MIP-1alpha and MIP-1beta, which promote activation of macrophages and NK cells, alongside increased IFN-gamma and TNF-alpha concentrations ^20^. Viral load also correlated significantly with IP-10 ^20^. In addition, tPA levels have been shown to increase in both HCPS and HFRS, whereas PAI-1 correlated with disease severity in HCPS but not in HFRS ^21^.

Identifying the host response factors that correlate with clinical outcome is therefore important for monitoring disease progression and identifying future therapeutic interventions. Prognostic biomarkers could support early risk stratification, guide monitoring intensity, and identify patients at risk of severe pulmonary, renal, or circulatory complications. At the same time, outcome-associated biomarkers may help define the host-response pathways that distinguish controlled infection from severe immunopathology. In this context, the present study characterises the host response with clinical outcomes in orthohantavirus infection, with the aim of providing further insight into the host and immune mechanisms that contribute to hantavirus disease severity.

## Results

Several different approaches can characterise a molecular phenotype to associate with disease severity, including transcriptomics and/or proteomics of clinical tissue. For example, transcriptomic analyses of blood samples taken from patients with Ebola virus disease (EVD) have been used to characterize disease severity and identify biomarkers in acutely ill patients that correlate with subsequent outcome (survival/death) ^22^. Single or multi ‘omic analysis of plasma samples have also been used to identify biomarkers that correlate with clinical outcomes, including in EVD ^23, 24^. To identify factors that may correlate with disease severity and HFRS, plasma samples from positively diagnosed patients were analyzed using a label free quantitative proteomics approach. To enhance broader applicability, longitudinal samples were obtained from patients infected with either DOBV or PUUV (Figure 2), and whose initial infection had been confirmed with RT-PCR (Table 1).

**Figure 2.**
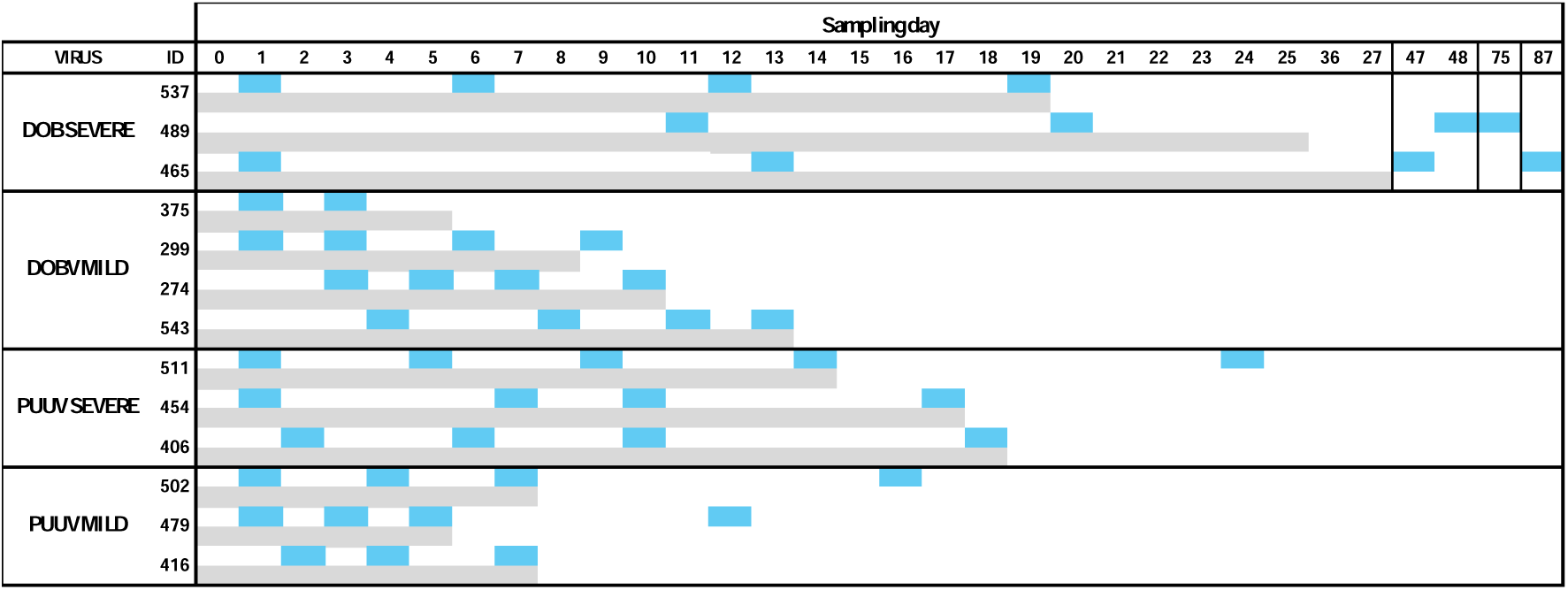
Sampling timeline (day post admittance) of patients with DOBV or PUUV and classified into having a severe or mild infection. The hospitalization period is shown in grey and the day of sample collection in blue.

**Table 1.** Details of patients with either DOBV, PUUV, or controls included in this study. Table shows sample number, virus identified, clinical severity, patient ID, day post-hospitalization the sample was taken, whether the patient was RT-PCR positive (+) or negative (-ve) for virus when the sample was taken, stage of infection and major clinical symptoms.

| No. | VIRUS | SEVERITY | Patient ID | Hospitalization (day) | Viral load, RNA copies/ml | Stage | MAJOR CLINICAL SYMPTOMS AND NOTES |
| --- | --- | --- | --- | --- | --- | --- | --- |
| 1 | DOBV | SEVERE | 537 | 1 | 411432 | Early | Blood transfusion, dialysis, acute kidney injury (AKI), pleural effusion, thrombocytopenia. |
| 2 |  |  |  | 6 | 37872 | Mid |  |
| 3 |  |  |  | 12 | 23732 | Mid |  |
| 4 |  |  |  | 19 | 15451 | Late |  |
|  | DOBV | SEVERE | 498 | 2 | 107544 |  | Viral load measured but no sample taken for proteomics. |
| 5 |  |  |  | 11 | 60525 | Mid | Blood transfusion, dialysis, AKI, pleural effusion, thrombocytopenia. |
| 6 |  |  |  | 20 | 7758 | Late |  |
| 7 |  |  |  | 48 | 0 | Late |  |
| 8 |  |  |  | 75 | 0 | Late |  |
| 9 | DOBV | SEVERE | 465 | 1 | 304500 | Early | Dialysis, AKI, bleeding, pleural effusion, thrombocytopenia. |
| 10 |  |  |  | 13 | 1475 | Mid |  |
| 11 |  |  |  | 47 | 0 | Late |  |
| 12 |  |  |  | 87 | 0 | Late |  |
| 13 | DOBV | MILD | 375 | 1 | 5543 | Early | Pleural effusion, vision blurriness. |
| 14 |  |  |  | 3 | 2140 | Early |  |
| 15 | DOBV | MILD | 299 | 1 | 34510 | Early | AKI, increased creatinine and AST/ALT levels, thrombocytopenia. |
| 16 |  |  |  | 3 | 68950 | Early |  |
| 17 |  |  |  | 6 | 2408 | Mid |  |
| 18 |  |  |  | 9 | 196 | Mid |  |
| 19 | DOBV | MILD | 274 | 3 | 39550 | Early | AKI, increased creatinine. |
| 20 |  |  |  | 5 | 26480 | Mid |  |
| 21 |  |  |  | 7 | 10240 | Mid |  |
| 22 |  |  |  | 10 | 6800 | Mid |  |
| 23 | DOBV | MILD | 543 | 4 | 160934 | Early | AKI, increased creatinine, vomiting. |
| 24 |  |  |  | 8 | 48100 | Mid |  |
| 25 |  |  |  | 11 | 26972 | Mid |  |
| 26 |  |  |  | 13 | 32116 | Mid |  |
| 27 | PUUV | SEVERE | 511 | 1 | 14722 | Early | Bleeding, thrombocytopenia. |
| 28 |  |  |  | 5 | 9658 | Mid |  |
| 29 |  |  |  | 9 | 414 | Mid |  |
| 30 |  |  |  | 14 | 0 | Mid |  |
| 31 |  |  |  | 24 | 0 | Late |  |
| 32 | PUUV | SEVERE | 454 | 1 | 55491 | Early | AKI, dialysis, thrombocytopenia. |
| 33 |  |  |  | 7 | 6792 | Mid |  |
| 34 |  |  |  | 10 | 0 | Mid |  |
| 35 |  |  |  | 17 | 0 | Late |  |
| 36 | PUUV | SEVERE | 406 | 2 | 58145 | Early | Bleeding, thrombocytopenia. |
| 37 |  |  |  | 6 | 5250 | Mid |  |
| 38 |  |  |  | 10 | 80 | Mid |  |
| 39 |  |  |  | 18 | 0 | Late |  |
| 40 | PUUV | MILD | 502 | 1 | 9567 | Early | Fever >38°C. |
| 41 |  |  |  | 4 | 245 | Early |  |
| 42 |  |  |  | 7 | 0 | Mid |  |
| 43 |  |  |  | 16 | 0 | Late |  |
| 44 | PUUV | MILD | 479 | 1 | 25212 | Early | Fever >38°C. |
| 45 |  |  |  | 3 | 13412 | Early |  |
| 46 |  |  |  | 5 | 858 | Mid |  |
| 47 |  |  |  | 12 | 50 | Mid |  |
| 48 | PUUV | MILD | 416 | 2 | 13823 | Early | Increased creatinine and<br>AST/ALT levels. |
| 49 |  |  |  | 4 | 3860 | Early |  |
| 50 |  |  |  | 7 | 41 | Mid |  |
| 51 | HEALTHY CONTROL 1 |  |  |  |  |  |  |
| 52 | HEALTHY CONTROL 2 |  |  |  |  |  |  |
| 53 | HEALTHY CONTROL 3 |  |  |  |  |  |  |
| 54 | HEALTHY CONTROL 4 |  |  |  |  |  |  |
| 55 | HEALTHY CONTROL 5 |  |  |  |  |  |  |

Patients were divided into those that had either mild or severe infection. The data indicated that patients with severe disease had longer hospital stays than patients with mild disease. In general patients with severe DOBV had the longest stays and patients with mild PUUV had the shortest (Figure 2). Viral load measurement on the first sample taken per patient (range 1 to 11 days after admission) indicated that on average patients who went on to have severe DOBV had the numerically higher average load, followed by patients who mild DOBV, severe PUUV and mild PUUV (average 274,492; 60,134; 42,786 and 16,200 RNA/copies ml respectively). Viral load declined over time within patients, and a linear mixed-effects model, suggested that PUUV showed a steeper decline than DOBV.

To characterise HFRS in patients infected with DOBV or PUUV, quantitative proteomic analysis was performed on plasma samples from mild and severe patients for either DOBV or PUUV across time points and compared to the negative controls. No sample depletion for high abundance proteins was used, and no late time point samples were analysed from mild individuals. A total of 589 proteins were identified, 361 of which were quantified across the samples after robust filtering (present in at least 50% of samples globally). From these, 175 proteins were found to be significantly different in abundance (adjusted p -value <0.05, >1 log_2_ fold abundance) between the sample groups and non-infected controls. The adjusted p-value controlled for multiple hypothesis testing, while the fold-change threshold ensured that identified proteins showed biologically meaningful abundance differences exceeding expected technical variability in label-free DDA quantification.

Principal component analysis (PCA) was used as an unsupervised approach to analyse and compare protein abundance values on a per patient/sample basis (Figure 3). The data indicated that samples from patients with either DOBV, PUUV, or the combined infected group clustered separately from the control group, independent of whether the disease was classified as mild or severe and even when virus had been cleared (as determined by a negative RT-PCR result). Interestingly, the trajectory of features from patient samples in PCA space showed a progressive shift from early to late time points toward the control cluster, consistent with partial resolution of the infection-associated proteomic signature over time.

**Figure 3.**
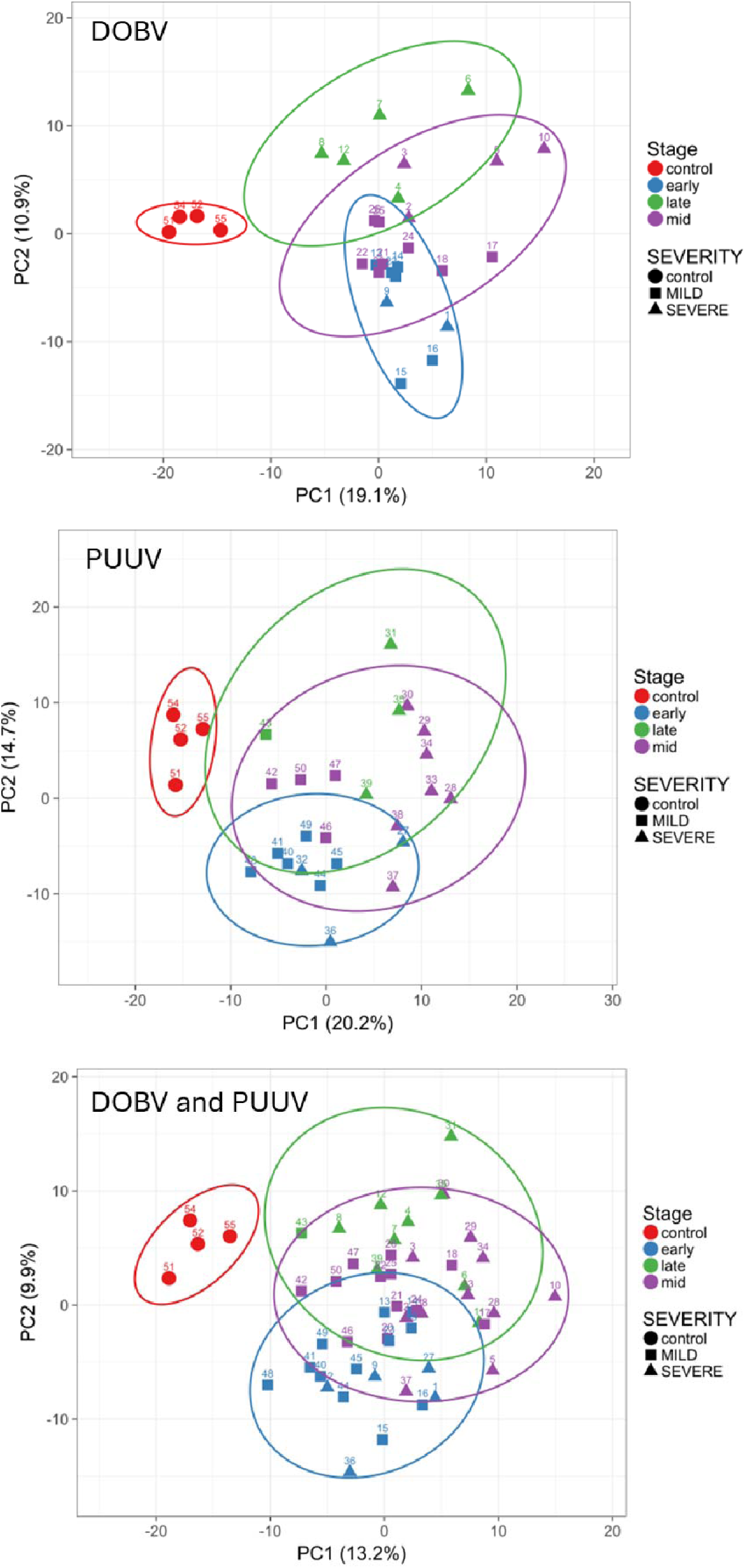
PCA of longitudinal proteomic profiles in DOBV and PUUV infection. Principal component analysis of proteomic data from longitudinal samples of patients infected with DOBV or PUUV and seronegative controls. Samples are coloured by infection stage (control, early with detectable viral RNA, mid, and late after viral clearance) and shaped by disease severity (control, mild, severe). Early-stage samples correspond to high viral load, whereas late-stage samples are defined by undetectable viral RNA by RT-PCR. The first two principal components (PC1 and PC2) are shown with the indicated variance explained. DOBV and PUUV cohorts are shown separately and combined. Sample 11 (DOBV) was excluded as an outlier due to its disproportionate influence on the visualisation of group separation.

The proteomic data was then analysed according to virus infection (DOBV or PUUV), with samples separated into early, middle and late time points (Figures 4 and 5). Irrespective of virus, disease severity or sampling time point, fibrinogen alpha, beta and gamma chains (FGA, FGB and FGG) exhibited the greatest fold compared with controls (Figures 4 and 5). Levels of C-reactive protein (CRP) also increased in early infection, decreasing at later time points, with fold change in abundance higher with PUUV compared with DOBV. Both fibrinogen and CRP are considered acute phase proteins whose circulating levels generally increase in response to inflammation ^25, 26^, as can arise during (or post) infection. At mid and late time points, elevated levels of immunoglobulins were observed for both viruses compared to the control group, including the V region of the variable domain of immunoglobulin light chains that are involved in antigen recognition (e.g. IGLV3-1, IGLV2-23, IGLV1-44).

**Figure 4.**
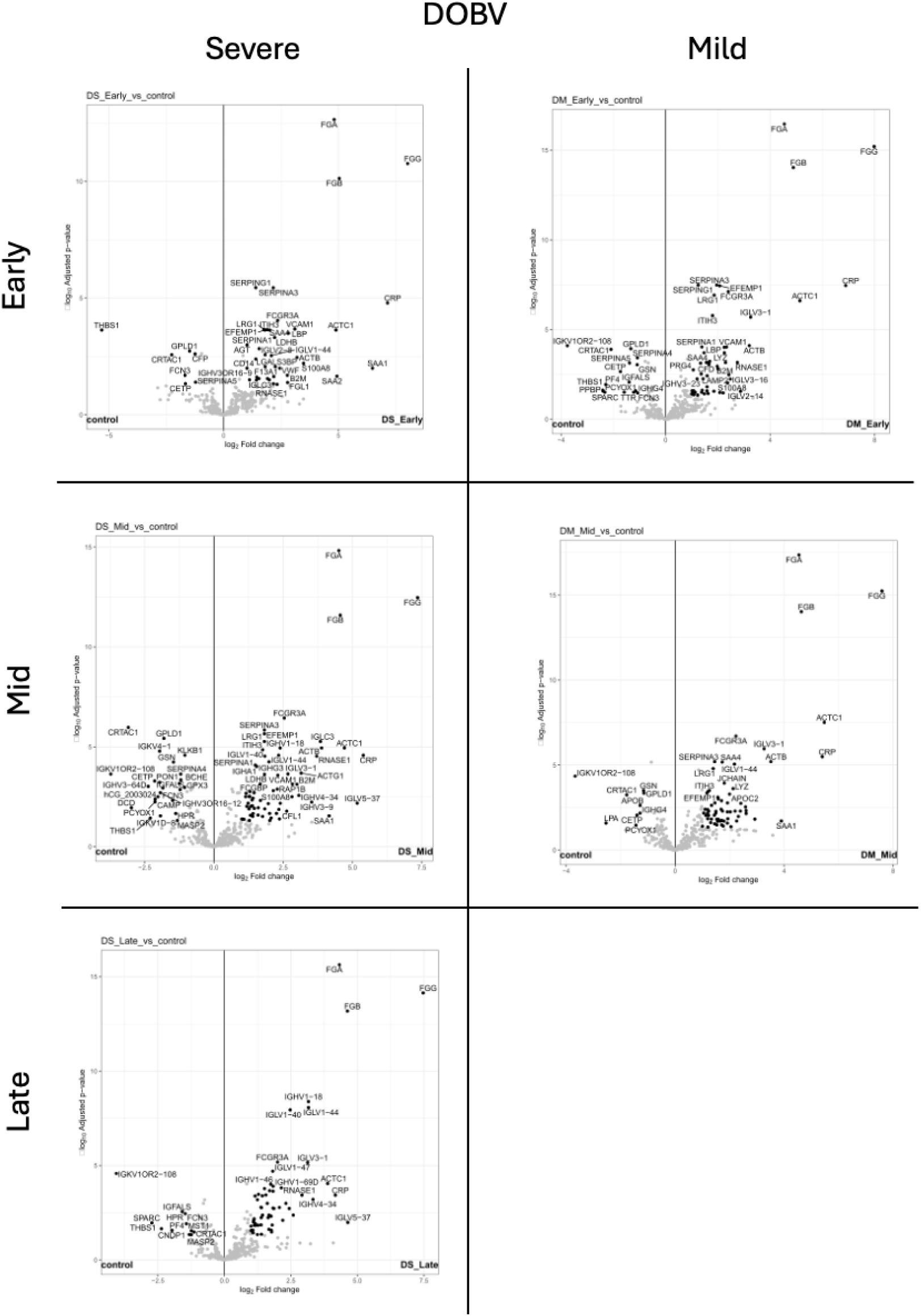
Volcano plots of protein abundance differences in plasma between patients infected with DOBV separated by time point (early, mid and late) and disease severity (severe and mild). Note there were no late samples from patients with mild disease.

**Figure 5.**
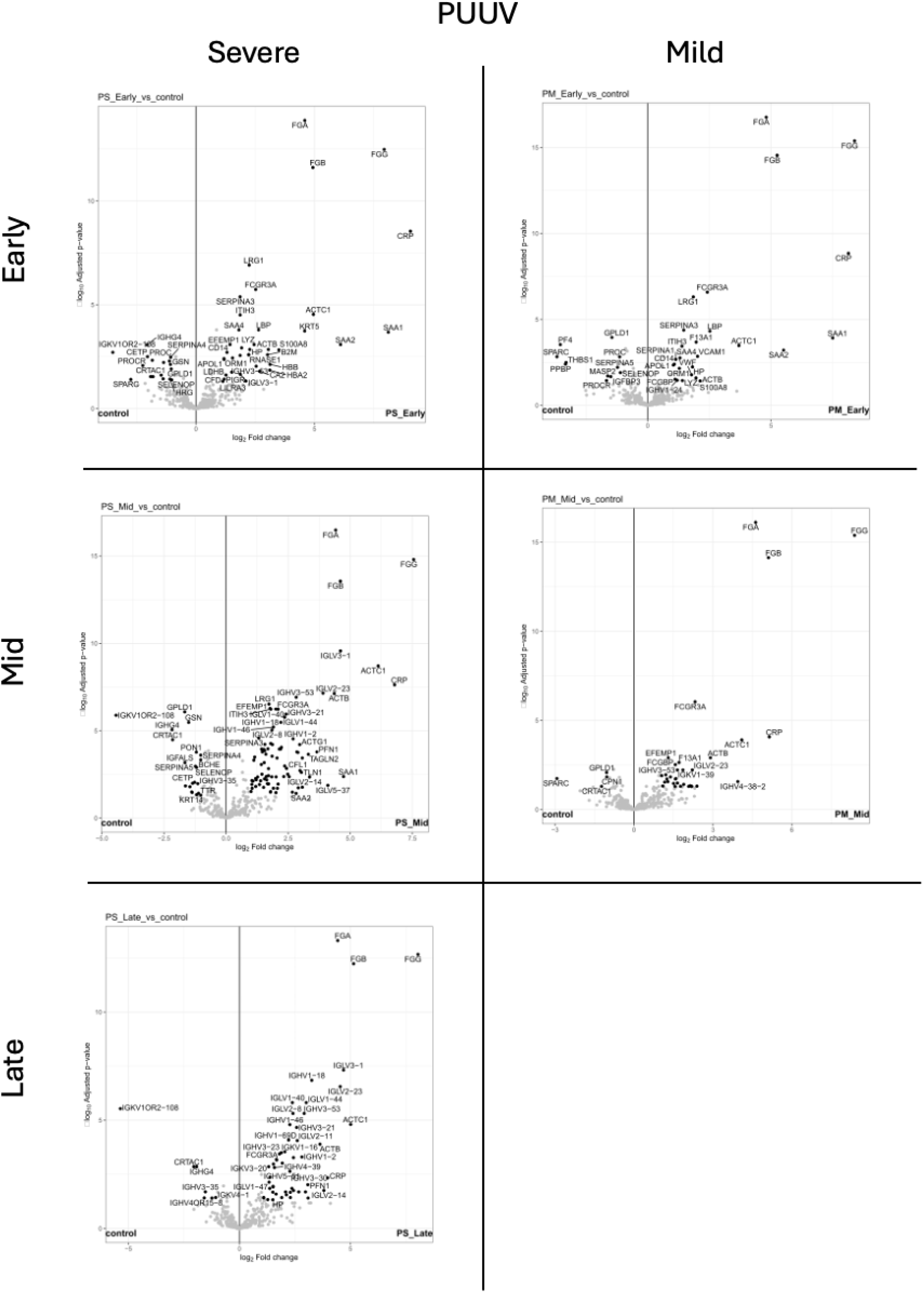
Volcano plots of protein abundance differences in plasma between patients infected with PUUV separated by time point (early, mid and late) and disease severity (severe and mild). Note there were no late samples from patients with mild disease.

To evaluate individual proteome levels changes in more detail, hierarchical clustering was performed for individual DOBV patient samples with mild and severe disease at early time points compared to control samples (Figure 6). DOBV-infected samples clearly differentiated from uninfected controls, with the overall protein abundance profiles sufficient to distinguish DOBV infection from no disease, regardless of disease severity (in agreement with the PCA analysis, Figure 3). Consistent regions of differential protein abundance were observed between mild and severe DOBV cases, with two primary clusters being identified (Figure 6).

**Figure 6.**
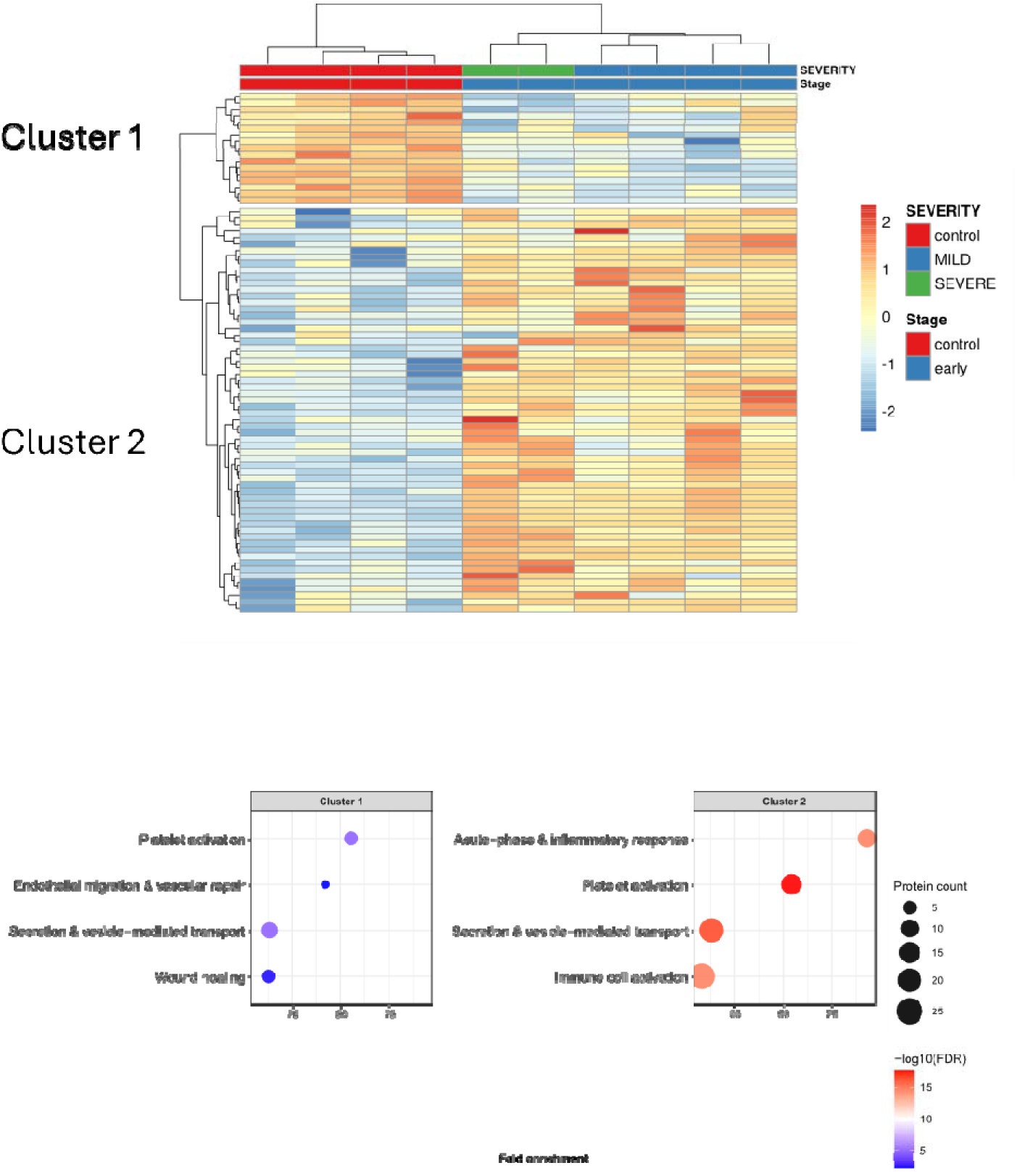
Early-stage DOBV infection is associated with distinct proteomic and functional signatures. Top: Heatmap of z-scored protein abundance with hierarchical clustering of samples and proteins from controls and patients with early-stage DOBV infection, annotated by disease severity. Two protein clusters show opposite expression patterns between control and disease groups. Bottom: Dot plots of enriched biological processes for each cluster. Dot size represents protein count and colour indicates −log10(FDR). Cluster 1 is enriched for vascular repair, platelet activation, and secretion pathways, whereas Cluster 2 is enriched for acute-phase, inflammatory, and immune responses.

Cluster 1 contained proteins lower in abundance in DOBV cases relative to controls, including platelet alpha-granule proteins (PF4 and PPBP) and thrombospondin-1 (THBS1), a platelet-derived matricellular protein with roles in coagulation and endothelial regulation. Complement-associated proteins such as ficolin-3 (FCN3) and complement factor properdin (CFP) were also reduced, consistent with active complement engagement and depletion upon infection [27]. Other depleted proteins included the negative acute-phase protein transthyretin (TTR), gelsolin (GSN, known to be reduced in sepsis and other sever inflammatory states), and the protease inhibitors SERPINA4 and SERPINA5.

Cluster 2 encompassed proteins elevated in DOBV-infected patients and was broadly divisible into two sub-groups. The first sub-cluster (upper portion) was dominated by immunoglobulin variable- and constant-region proteins, including multiple IgHV and IgLV segments, IgM heavy chain (IGHM), and J-chain (JCHAIN), consistent with an active primary or ongoing humoral response at these early disease time points. FCGR3A (CD16a), RNASE1, and LYZ were co-elevated with this immunoglobulin group, pointing to concurrent NK cell and neutrophil activation alongside the humoral response. Fibronectin (FN1) was also elevated, consistent with extracellular matrix re-modelling and inflammatory cell recruitment.

The lower portion of Cluster 2 contained acute-phase, inflammatory, endothelial, and coagulation-associated proteins, many of which were preferentially elevated in severe cases. The acute-phase associated proteins FGL1 (fibrinogen-like protein 1), SAA1 and SAA2 were elevated in the more severe patients. Of particular interest was the concurrent elevation of these with CRP, LBP, CD14, (ORM1) and S100A8, indicating coordinated activation of innate immune pathways, linking LPS recognition (through the LBP/CD14/TLR4 axis) with downstream myeloid cell activation and amplification of inflammation via S100A8 in this systemic acute-phase response. More broadly, this cluster included FGA, FGB FGG, and a key acute-phase protein involved in coagulation and immunothrombosis, endothelial activation markers (VCAM1, ICAM1), the complement pathway regulator SERPING1 (C1-inhibitor), and multiple immunoglobulin variable/constant region proteins.

Clustering of early time-point samples of PUUV-infected patients compared to controls, showed a clear infection-associated proteomic signature (Figure 7). As with DOBV, control samples clustered separately from infected samples, and severity differences were most apparent within individual protein clusters rather than at the level of global sample topology.

**Figure 7.**
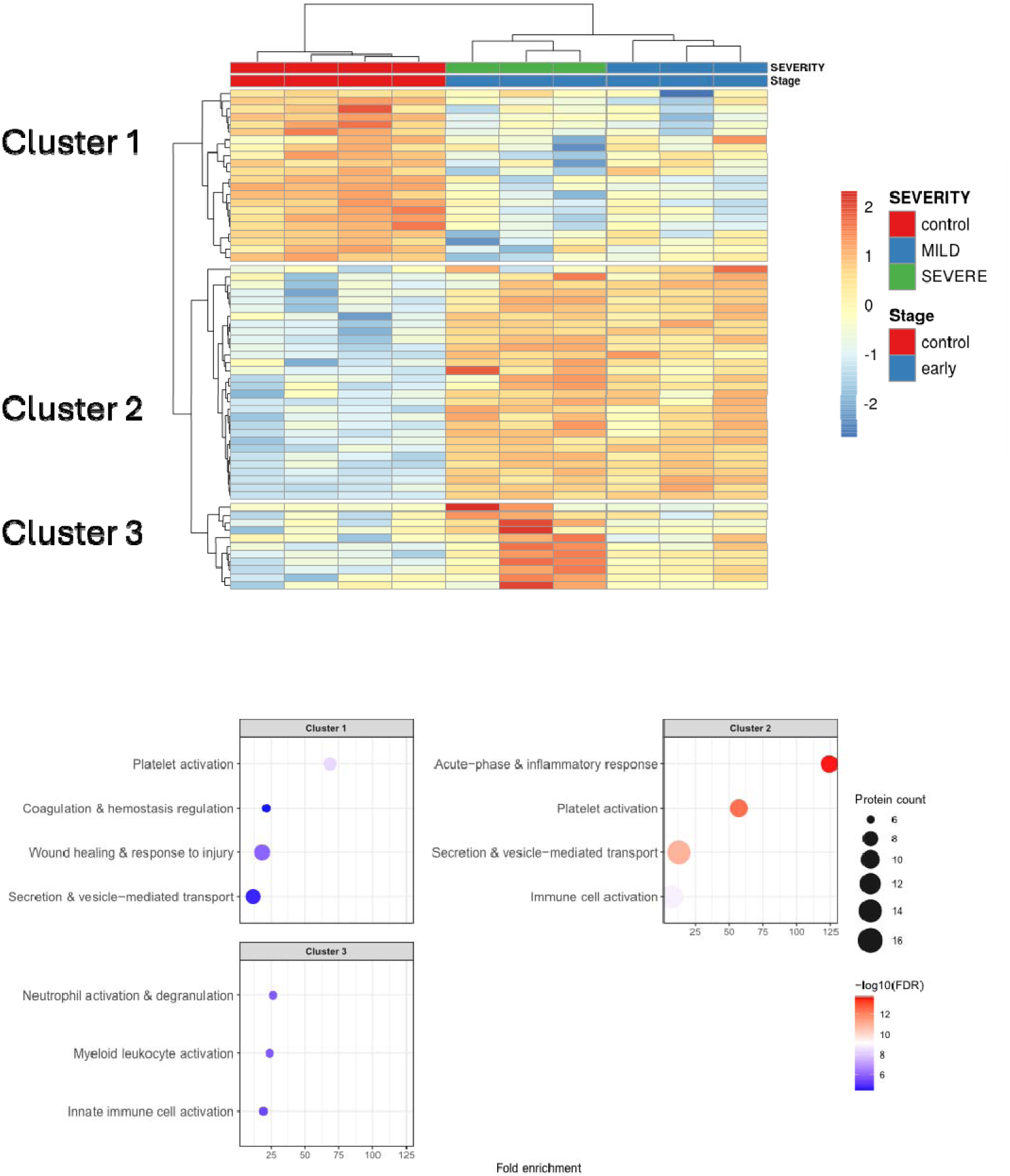
Early-stage PUUV infection is associated with distinct proteomic and functional signatures. Top: Heatmap of z-scored protein abundance with hierarchical clustering of samples and proteins from controls and patients with early-stage PUUV infection, annotated by disease severity. Three protein clusters show distinct expression patterns across groups. Bottom: Dot plots of enriched biological processes for each cluster. Dot size represents protein count and colour indicates −log10(FDR). Cluster 1 is enriched for platelet activation, coagulation, and wound-healing pathways; Cluster 2 for acute-phase, inflammatory, and immune responses; and Cluster 3 for neutrophil and innate immune activation.

Cluster 1 contained proteins depleted in PUUV-infected patients. Like samples from DOBV infected patients, PF4, PPBP, and THBS1 were reduced, with more pronounced depletion in severe cases. GSN, SPARC, SERPINA4, and TTR were similarly reduced, recapitulating the DOBV pattern. The coordinated depletion of platelet granule proteins and anticoagulant pathway components in Cluster 1 is consistent with the platelet activation, thrombocytopenia, and coagulation disturbance that are clinical hallmarks of HFRS [28]. In contrast to DOBV samples, the anticoagulant proteins Protein C (PROC) and its endothelial receptor (PROCR) were depleted in PUUV-infected patients and to a greater degree in severe cases, indicating downregulation of the Protein C pathway. Interestingly, DOBV samples were depleted in levels of the PKC inhibitor SERPINA5, with a marked reduction particularly in severe cases, suggesting contrary regulation of the PKC pathway in these two infection systems.

Cluster 2 contained proteins elevated in PUUV-infected patients relative to controls, with the strongest signals in severe cases. Consistent with the DOBV dataset, a broad acute-phase response signature was observed, comprising SAA1, SAA2, and SAA4 alongside CRP, ORM1, LBP, CD14, SERPINA1, SERPINA3, and ITIH3. Endothelial activation was evidenced by elevated VWF, and LGALS3BP levels were again increased in severe cases, consistent with its identification as a cross-virus severity-associated candidate. Further proteins elevated in infected patients and consistent with DOBV findings included LRG1, implicated in endothelial dysfunction through TGF-β signaling; F13A1, a fibrin-crosslinking coagulation factor; APOL1, an innate immune effector; and LILRA3, an inhibitory receptor expressed on monocytes and NK cells.

Cluster 3 was the most severity-specific cluster, comprising proteins near-baseline in controls and mild cases but consistently elevated across severe PUUV patients. S100A8, B2M, LYZ, and RNASE1 were co-elevated in severe cases; the combination of a neutrophil granule protein (LYZ), a secreted ribonuclease (RNASE1), and a DAMP alarmin (S100A8) in severe but not mild disease is consistent with neutrophil degranulation and possible NET formation, contributing to the severe disease phenotype. CFD (complement factor D), the rate-limiting enzyme of the alternative complement pathway, was elevated specifically in two of the three severe cases, providing direct proteomic support for alternative pathway activation in severe PUUV infection [27]. LAMP2 elevation across all infected samples may reflect lysosomal exocytosis from activated immune or endothelial cells.

### Early severity effect-size analysis

To test whether changes in protein abundance in the early phase were associated with later severity, early samples were collapsed within each patient to define the median relative abundance for each protein. For each protein p, the Δp = median_s∈severe_(M_p,s_) − median_s∈mild_(m_p,s_) was computed, where m_p,s_ is the early subject-level median abundance. Positive Δp values indicated higher early abundance in severe cases; negative values indicated lower abundance. Exact subject-label permutation tests were used, but the small number of subjects made the p-values discrete and low-resolution. Results are therefore interpreted as effect-size rankings for candidate prioritisation rather than formal hypothesis testing.

In the combined DOBV+PUUV early analysis, 14 proteins had raw exact permutation p < 0.05, but none survived FDR correction across all 361 identified proteins (Table 2). Virus-specific analyses were more limited: DOBV had only 15 exact permutations (minimum achievable p-value = 0.0667) and PUUV had 20 (minimum p-value = 0.05), precluding formal significance testing. Virus-specific results in Table 3 were therefore interpreted as effect-size rankings only.

**Table 2:**
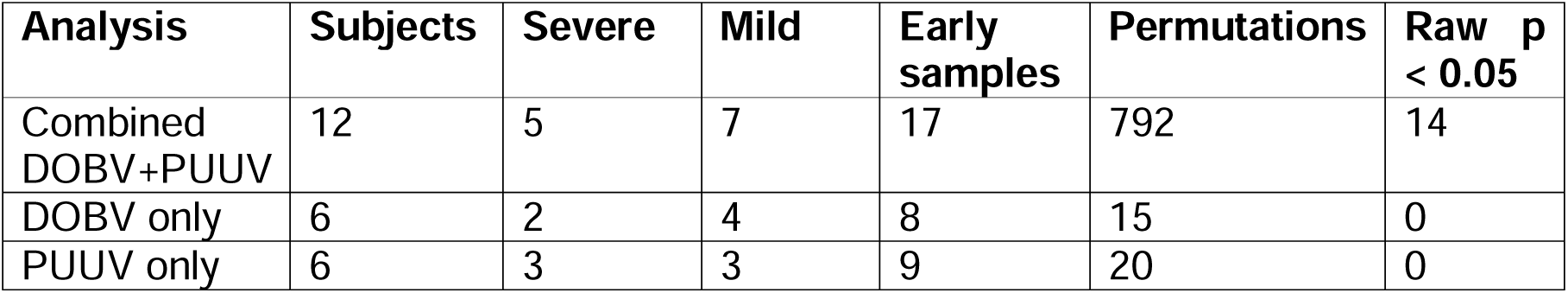
Early severity analysis summary.

| Analysis | Subjects | Severe | Mild | Early samples | Permutations | Raw $p < 0.05$ |
| --- | --- | --- | --- | --- | --- | --- |
| Combined DOBV+PUUV | 12 | 5 | 7 | 17 | 792 | 14 |
| DOBV only | 6 | 2 | 4 | 8 | 15 | 0 |
| PUUV only | 6 | 3 | 3 | 9 | 20 | 0 |

**Table 3:** Top candidates from the combined early severity analysis.

| Protein | Description | $\Delta_p$ severe-mild | Exact p |
| --- | --- | --- | --- |
| IGKV2-28 | Immunoglobulin kappa variable 2-28 | 2.107 | 0.0126 |
| LGALS3BP | Galectin-3-binding protein | 1.083 | 0.0126 |
| APOH | Beta-2-glycoprotein 1 | -0.637 | 0.0126 |
| S100A9 | Protein S100-A9 | 1.625 | 0.0253 |
| IGHG1 | Immunoglobulin heavy constant gamma 1 | -0.423 | 0.0253 |
| CPN2 | Carboxypeptidase N subunit 2 | 0.419 | 0.0253 |
| C4BPB | C4b-binding protein beta chain | 0.314 | 0.0253 |
| S100A8 | Protein S100-A8 | 1.523 | 0.0328 |
| CAMP | Cathelicidin antimicrobial peptide | 1.482 | 0.0328 |
| PRDX2 | Peroxiredoxin-2 | 1.168 | 0.0328 |
| SERPINA10 | Protein Z-dependent protease inhibitor | 0.503 | 0.0328 |
| IGHV1-8 | Immunoglobulin heavy variable 1-8 | 3.916 | 0.0328 |

The strongest early severity candidates in the combined analysis included S100A8, S100A9, CAMP, LGALS3BP, and PRDX2, together with the complement and coagulation associated proteins CPN2, C4BPB, SERPINA10, and APOH (Table 3).

The elevation of S100A8 (Δ_p_ = 1.523) and S100A9 (Δ_p_ = 1.625) in severe cases is consistent with their appearance as severity-associated proteins in the hierarchical clustering of proteins in early-stage samples from DOBV and PUUV (Figure x), where they were among the most visually prominent severity-specific signals, particularly in PUUV Cluster 3. LGALS3BP (Δ_p_ = 1.083) also appeared as a severity-associated protein in Cluster 2 for both viruses, making it the best-supported cross-virus severity candidate.

### Changes in the abundance of some host proteins were virus specific

Virus-specific early rankings were underpowered for formal inference but informative for candidate selection. In patients with DOBV, large severe–mild effects were observed for IGHV1-8, IGHV5-10-1, THBS1, SAA1, SAA2, FGL1 and ACTN1. The effect size rankings for SAA1, SAA2 and THBS1 are consistent with the DOBV heatmap, where SAA1 and SAA2 showed the strongest severity-associated signals in Cluster 2 and THBS1 was more depleted in severe than mild cases in Cluster 1. In patients with PUUV, the largest effect-size rankings included B2M, LPA, ICAM2, HBA2, IGKV2-28, PPIA, and LYZ. The elevation of B2M and LYZ in severe PUUV corroborates their appearance in PUUV Cluster 3, where both were among the proteins most elevated in severe cases, identifying them as exploratory candidates for further investigation.

Overall, the early severity analysis identified proteomic variation with clear infected–control separation and severity-associated signals, particularly in PUUV patients. The convergence of top-ranked candidates, across both the effect-size rankings and the heatmap clustering analyses, specifically S100A8, S100A9, LGALS3BP, SAA1, SAA2, and B2M, strengthens confidence in these proteins as the most robustly supported early severity candidates in the dataset, despite the small sample size and the absence of FDR-corrected significance. Cleaner trajectory summaries for these candidate proteins were subsequently generated to examine longitudinal behaviour across disease course.

Individual sample values were plotted as points, while group-level lines show the median abundance within each virus–severity–stage group. This avoided overinterpreting irregular individual sampling while retaining the longitudinal structure.

Figure 8 summarizes the trajectory patterns for the main candidate proteins, with positive values indicated higher abundance in severe cases, and negative values indicated lower abundance. Late-stage effects in patients with DOBV are not shown as the sampling structure was too sparse for a reliable severe–mild stage contrast at that time point.

**Figure 8:**
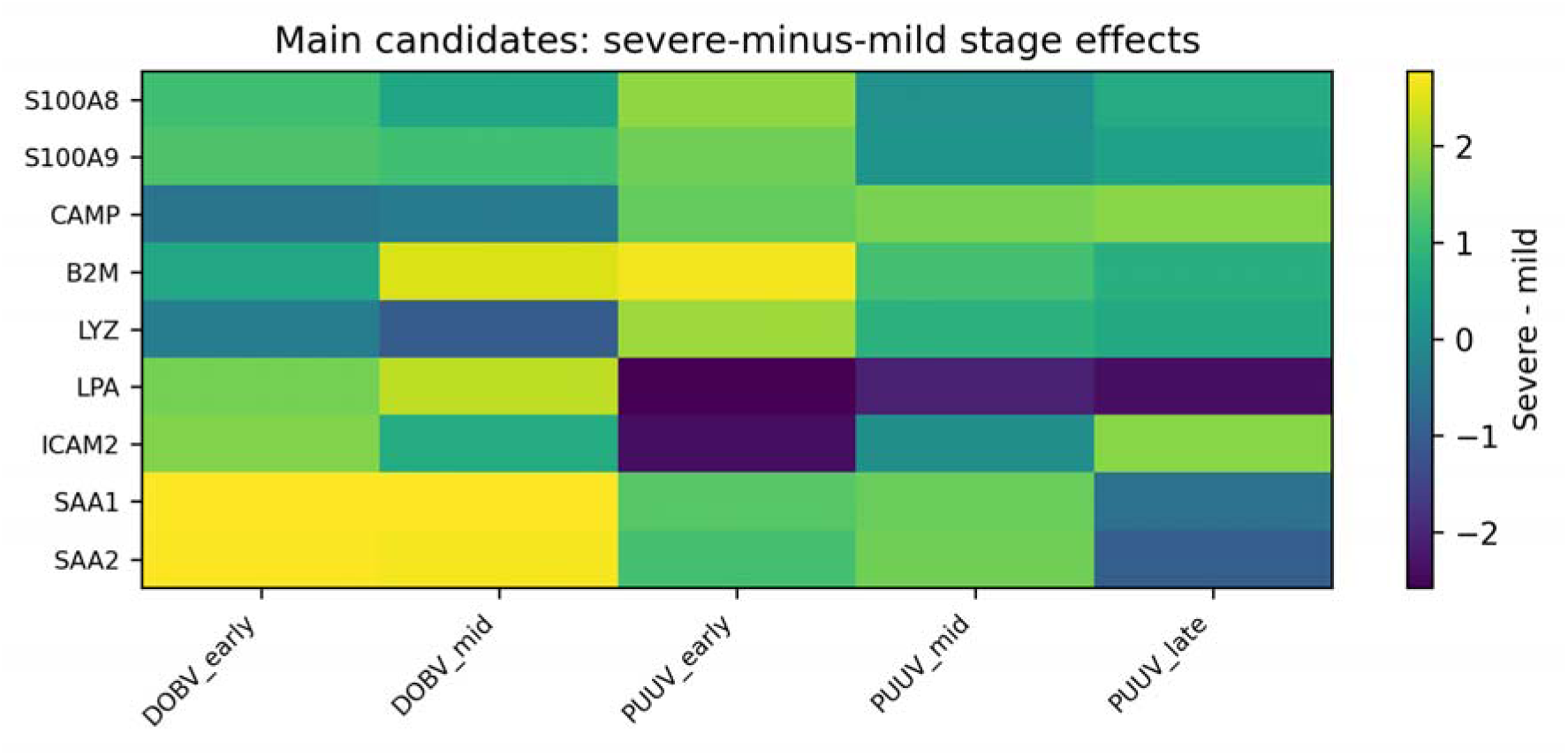
Severe-minus-mild stage effects for main candidate proteins.

The clearest and most consistent severity-associated patterns were observed for SAA1, SAA2, B2M, S100A8, and S100A9. SAA1 and SAA2 were substantially higher in severe cases at early- and mid- time points in both DOBV and PUUV, consistent with their prominence as severity-associated proteins in the early heatmap clustering, and with their identification as top candidates in the early effect-size analysis. S100A8 and S100A9 were elevated in severe cases at early time-points across both viruses, with the effect attenuating at later stages. This temporal pattern is consistent with an early innate immune amplification signal, rather than a sustained one. It also corroborates their appearance as severity-specific proteins in PUUV Cluster 3 and the combined early severity rankings (S100A8 Δ = 1.523, S100A9 Δ = 1.625, both p = 0.0328). B2M was higher in severe cases particularly at early time points in PUUV and mid time points in DOBV, in keeping with its detection as a severity-associated protein in the PUUV Cluster 3 analysis and in the longitudinal GEE models.

Several proteins showed virus-dependent severity behavior (Table 4). CAMP was higher in severe PUUV across stages but did not show the same pattern in DOBV. LYZ was higher in severe PUUV (as seen in PUUV Cluster 3), but lower in severe DOBV, representing a directional reversal between viral species. LPA and ICAM2 similarly showed opposite early-stage directions between DOBV and PUUV, indicating that not all severity-associated proteomic signals are conserved across hantavirus strains (Table 4).

**Table 4.** Main trajectory patterns among selected proteins.

| Pattern | Proteins | Interpretation |
| --- | --- | --- |
| Higher in severe, broadly consistent | SAA1, SAA2, B2M | Strongest severity-associated candidates |
| Higher in severe early samples | S100A8, S100A9 | Early inflammatory signal |
| Higher in severe PUUV | CAMP, LYZ | PUUV-dominant severity pattern |
| Virus-dependent direction | LPA, ICAM2, LYZ | Possible strain-specific biology |
| Sparse late-stage evidence | Several proteins | Late-stage interpretation limited by sampling |

### Analysis of longitudinal immune trajectories

To examine whether severity groups exhibited different recovery trajectories, and whether the strongest individual protein signals clustered into coherent pathophysiological modules, a longitudinal analysis was performed using the filtered non-imputed protein matrix across all infected subjects (n = 13). Candidate proteins were grouped into five modules on the basis of biological function: acute-phase inflammation (SAA1, SAA2), neutrophil/innate immunity (S100A8, S100A9, CAMP, LYZ), coagulation/complement/plasma biology (APOH, SERPINA10, C4BPB), endothelial and vascular biology (THBS1, ICAM2), and interferon/antigen-presentation biology (B2M, HLA-A). Module scores were computed by standardising available detected proteins across infected samples and taking the within-module median.

Given the small cohort size (n = 13), unbalanced severity and virus strata, and irregular follow-up, longitudinal models were kept deliberately simple and are presented as exploratory, with emphasis on effect direction, biological coherence, and consistency across methods rather than formal inference. Mixed-effects models and GEE models clustered by patient were fitted as robustness checks. The main model included severity, virus, and day of hospitalization. A second exploratory model additionally included a severity-by-day interaction to assess whether recovery kinetics differed between severity groups. Patient-level recovery slopes were also computed as descriptive summaries for focal proteins.

At the modular level (Figure 9), the acute-phase module showed the most interpretable recovery-kinetics pattern: mild cases declined more strongly over time while severe cases remained more persistently elevated. This pattern of slower inflammatory resolution in severe disease extends the cross-sectional heatmap findings, where SAA1 and SAA2 were most strongly elevated in severe early samples, into the longitudinal domain, and suggests that severity in HFRS is characterised not only by greater initial inflammatory activation but by impaired resolution of that response.

**Figure 9.**
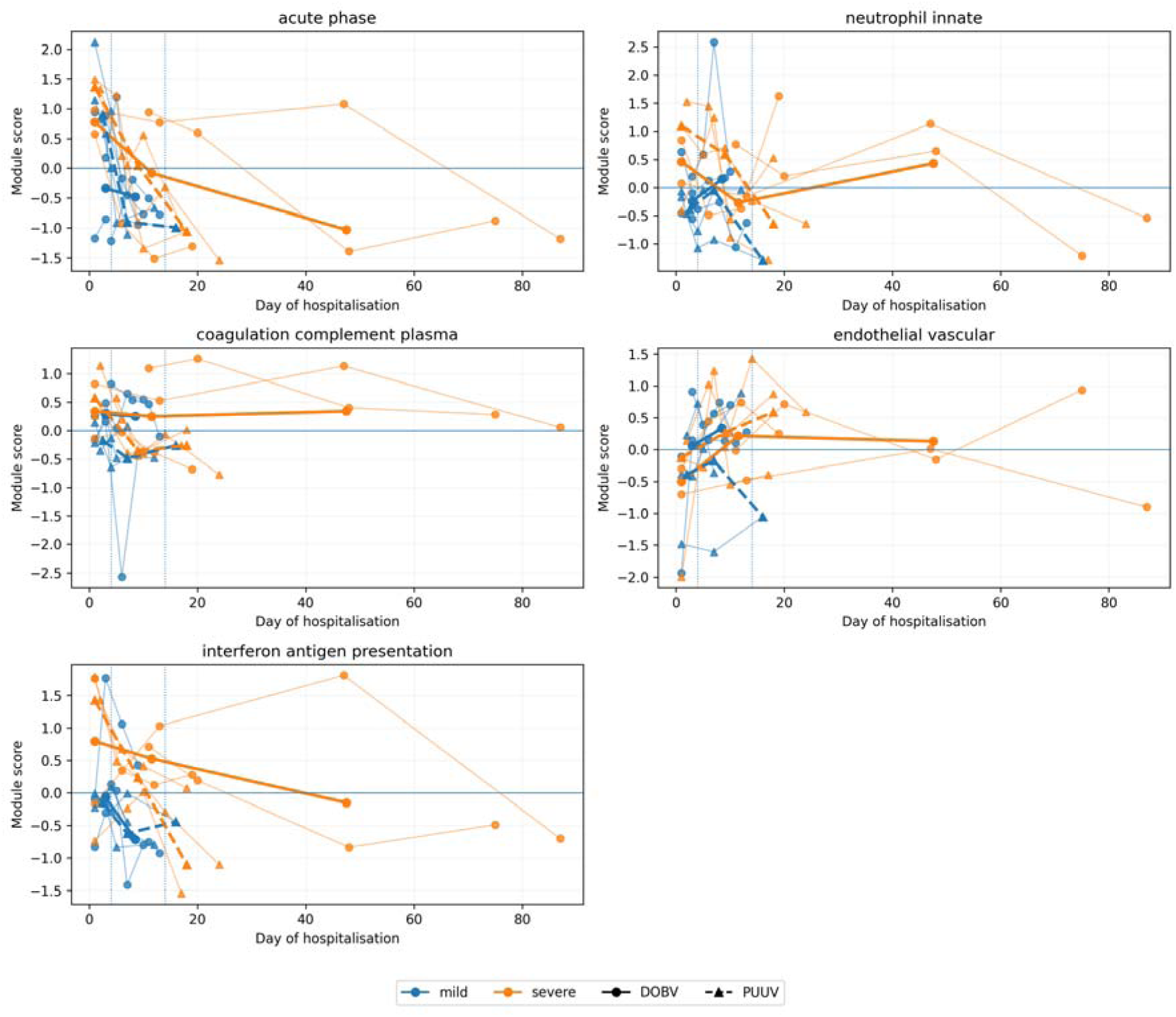
Pathophysiological module recovery trajectories using the filtered non-imputed matrix. Thin lines show individual patient trajectories and thick lines show group-level trends.

The neutrophil/innate immunity module was higher in severe cases at early and mid-time points, consistent with the early severity-specific elevation of S100A8 and S100A9 observed in the heatmap clustering and effect-size analysis, and with the biological role of these DAMP proteins in sustaining innate immune activation. The interferon/antigen-presentation module was also higher in severe disease, driven principally by B2M, whose severity-associated elevation was supported by GEE modelling at the individual protein level. The coagulation/complement/plasma module showed weaker but directionally consistent severe-associated elevation. The endothelial/vascular module was the least stable, driven in part by the sparse detection of ICAM2, and should be treated as exploratory.

At the individual protein level, GEE models provided supportive evidence for severe-associated elevation of B2M, S100A8, S100A9, LGALS3BP, and SERPINA10. Exploratory severity-by-day interaction terms were most consistent for SAA1, SAA2, B2M, LGALS3BP, and SERPINA10, suggesting that these proteins show not only higher abundance in severe disease, but also differential recovery kinetics. The convergence of LGALS3BP across the combined early effect-size analysis (Δ = 1.083, p = 0.0126), the heatmap clustering of both DOBV and PUUV, and the GEE longitudinal models makes it the most consistently supported cross-virus severity candidate in the dataset. Patient-level recovery slopes (Figure 10) showed substantial inter-subject heterogeneity. While the slope summaries for SAA1 and SAA2 show the clearest descriptive patterns, they were influenced by the availability of late follow-up concentrated in a small number of severe subjects.

**Figure 10:**
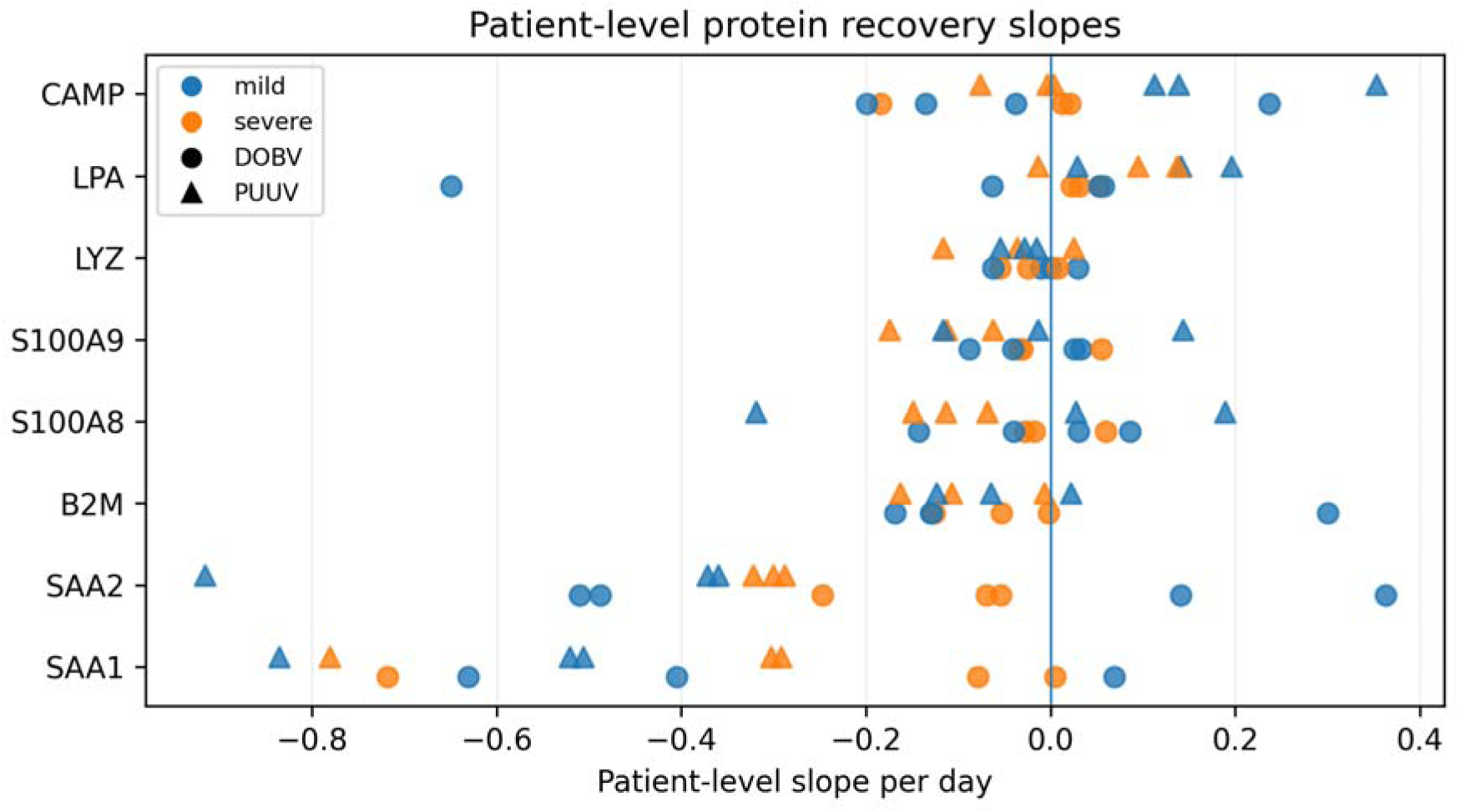
Patient-level recovery slopes for focal proteins. Each point represents one subject-level slope over observed follow up. Colour indicates severity and marker shape indicates virus.

Detection support was therefore reviewed alongside abundance trajectories. Core severity-associated proteins such as B2M, SAA2, S100A8 and S100A9 were detected in a high proportion of samples across virus and severity strata, supporting interpretation of their abundance trajectories. In contrast, CAMP, ICAM2 and SAA1 showed more condition- or stage-specific non-detection, particularly in later stages and in some milder disease strata. Module contributions were therefore treated as exploratory where detection was sparse.

## Discussion

This study provides a longitudinal plasma proteomic analysis of patients with haemorrhagic fever with renal syndrome caused by DOBV or PUUV, two clinically important Old-World hantaviruses associated with differing disease severity. We note that as well as being a single country cohort, this study is necessarily limited by the rarity of well-characterised longitudinal DOBV and PUUV cases, which occur sporadically. The samples were analysed without depletion of high-abundance plasma proteins, which preserved clinically abundant proteins such as fibrinogen and immunoglobulins but may have reduced sensitivity for lower-abundance cytokines, endothelial mediators, and tissue-injury markers. Low physiological levels of TNF-alpha through a specific genotype have been linked with a more severe disease in patients with PUUV ^27^. VEGF may have different effects depending on the stage of infection. For example, data suggests that during Hantaan virus infection, VEGF may be involved in renal disfunction during the oliguric stage but promote repair of renal injury during convalescence ^28^.

The data showed that hantavirus infection was associated with a persistent perturbation of the circulating proteome, with infected patients separating from healthy controls across early, mid, and late disease time points (Figure 3). This separation was still apparent after viral RNA was no longer detectable in some patients, suggesting that the systemic host response outlasted measurable viraemia and may have contributed to ongoing clinical pathology. This interpretation is consistent with current models of hantavirus disease, in which pathology reflects a combination of endothelial dysfunction, altered vascular permeability, platelet/coagulation abnormalities, complement activation, and immune-mediated inflammation rather than direct cytopathic destruction of infected endothelial cells ^3, 10^.

Despite the clinical differences between DOBV and PUUV infection, early-stage hierarchical clustering indicated a broadly shared host-response signature. Depletion of platelet alpha-granule proteins and baseline plasma proteins (Cluster 1), and elevation of acute-phase proteins, endothelial activation markers, innate immune, and severity-associated candidates aligned with proteolysis regulation, complement activation and platelet degranulation (Figure 5, 7) were common to both infections. This convergence supports a model in which DOBV and PUUV engage a shared set of host-response pathways, with severity reflecting quantitative amplification of those shared mechanisms rather than qualitatively distinct biology.

A major feature of the proteomic signature was the increased abundance of acute-phase proteins, particularly FGA, FGB, FGG and CRP, across both DOBV and PUUV infection (Figure 4). Fibrinogen and CRP are canonical systemic inflammatory proteins; CRP rises rapidly at sites of infection and inflammation, whereas fibrinogen has both haemostatic and immunomodulatory roles during tissue injury and inflammatory responses ^29, 30^. Increases in fibrinogen have been observed in patients in the acute phase of Ebola virus disease ^22^. The increase in abundance of these proteins in patients with HFRS compared with controls suggested a robust hepatic acute-phase response. Because fibrinogen and CRP were elevated in both mild and severe disease groups, they can be interpreted as markers of infection-associated inflammation rather than stand-alone discriminators of clinical severity.

The absence of a clear relationship between initial viral load and disease severity in this cohort is also informative. Although viral replication is required to initiate disease, severe HFRS may be determined less by viral load at hospital presentation than by downstream host-response pathways. This is consistent with the hypothesis that disease severity is frequently linked to immune activation, vascular leakage, coagulation imbalance, complement activation and platelet dysfunction. In PUUV infection specifically, host inflammatory mechanisms, endothelial barrier dysfunction, thrombocytopenia, and acute kidney injury have been recognised as central clinical and pathogenic features ^19^.

Complement activation has previously been demonstrated in acute PUUV infection and shown to correlate with disease severity, including clinical and laboratory markers of severe disease ^31^. Complement activation could contribute directly to endothelial activation and capillary leakage, but may also reflect immune-complex formation, tissue injury, or impaired regulation of innate immune effector pathways. The identification of complement-related proteins in the present proteomic dataset therefore supports the concept that complement dysregulation is a shared component of severe hantavirus disease, and not merely a secondary marker of inflammation.

The platelet-degranulation signature is also consistent with known clinical and mechanistic features of hantavirus infection. Thrombocytopenia is one of the defining laboratory abnormalities of HFRS ^32^, and previous work has shown that HFRS is accompanied by increased thrombopoiesis, platelet activation, reduced ex vivo platelet function, and evidence that platelet activation contributes to intravascular coagulation ^33^. In this context, differences in platelet-associated proteins between mild and severe infection may reflect platelet consumption, activation, degranulation, endothelial adhesion, or altered megakaryopoiesis. Because platelet dysfunction can intersect with vascular leakage, bleeding, and coagulation abnormalities, these proteins are plausible candidate biomarkers for risk stratification of disease severity in patients.

The increase in immunoglobulin variable-region proteins at mid and late time points indicates development of the adaptive humoral response during clinical progression and recovery. Although obvious, this is consistent with the importance of antibody responses in hantavirus infection, including evidence that humoral immunity contributes to recovery in hantavirus cardiopulmonary syndrome ^16^. The temporal pattern observed here suggests that longitudinal proteomics can capture the transition from an early innate/acute-phase response toward a later adaptive immune profile. Importantly, this may help distinguish markers of acute disease severity from markers of immune resolution or convalescence.

Interestingly, DOBV infection was associated with a substantially larger circulating immunoglobulin signal at early time points, suggesting more extensive or earlier humoral mobilisation relative to PUUV. Complement depletion in DOBV Cluster 1 was characterised by loss of the lectin pathway component FCN3 and the alternative pathway regulator CFP, whereas PUUV Cluster 1 showed more prominent depletion of the anticoagulant Protein C pathway (PROC, PROCR). Whether these differences reflect viral tropism, disease kinetics, or host genetic factors in the respective patient cohorts cannot be determined from this relatively limited analysis but identifies candidate pathway-level differences that warrant further investigation in larger cohorts.

The data also reinforce the central role of the endothelium in hantavirus disease. Pathogenic hantaviruses infect endothelial cells without causing overt lysis yet can sensitise endothelial barriers to permeability mediators such as VEGF ^13^. Hantavirus-infected cells may also resist cytotoxic lymphocyte-mediated apoptosis, providing a mechanism by which infected endothelial cells persist while remaining targets of immune activation ^11^. The proteomic enrichment of inflammatory, coagulation, complement, and platelet-associated pathways in the present study is therefore compatible with an endothelial-centred model in which vascular dysfunction is driven by host-response amplification rather than direct virus-induced cytopathology.

The comparison between patients infected with DOBV and patients infected with PUUV provided a robust analysis of HFRS, however both viruses have been reported to differ in typical clinical severity ^3, 7^. This can be seen from the data where patients infected with DOBV were associated with greater clinical severity and a longer stay time than patients infected with PUUV. Despite these clinical differences, the broad proteomic response observed here shared common features across both viruses, particularly acute-phase activation and perturbation of coagulation-related proteins. This suggests that during infection DOBV and PUUV may engage overlapping host-response pathways, while differences in magnitude, timing, tissue tropism, or host susceptibility may determine clinical outcome. The observation that LGALS3BP was shared among severity-associated proteins in both DOBV and PUUV patient groups is of interest as galectin-family proteins have been identified as markers of inflammation, endothelial activation, and tissue injury ^34^.

Taken together, the longitudinal analysis strengthens the biological interpretation of the first-pass findings (Table 6). The main signal is not simply a collection of isolated protein differences, but a coordinated pattern involving acute phase inflammation, innate/neutrophil activation, and interferon/antigen-presentation biology. The GEE robustness analysis was directionally consistent with higher neutrophil/innate and interferon/antigen-presentation module scores in severe disease, supporting the interpretation that severe HFRS is associated with stronger inflammatory and antiviral immune activation. A central observation is the apparent difference in inflammatory recovery kinetics between mild and severe disease. The acute-phase module showed a strong negative time effect overall, consistent with resolution of inflammation over time, together with a positive severity-by-day interaction, indicating slower decline in severe cases. This pattern was visible at the patient level for SAA1 and SAA2, where mild cases generally showed steeper declines while several subjects with severe disease retained elevated values into later follow-up. The patient-level recovery slopes were therefore most consistent with possible delayed or incomplete resolution of the acute-phase response in severe disease. Slow or incomplete resolution of acute-phase inflammation may be a clinically meaningful feature of severe HFRS and a candidate signal for identifying patients at risk of prolonged or complicated recovery. These findings support further investigation of early, targeted host-response interventions aimed at limiting inflammatory amplification, endothelial leakage and organ failure, while recognising the need to manage renal dysfunction and bleeding risk in HFRS.

**Table 6:**
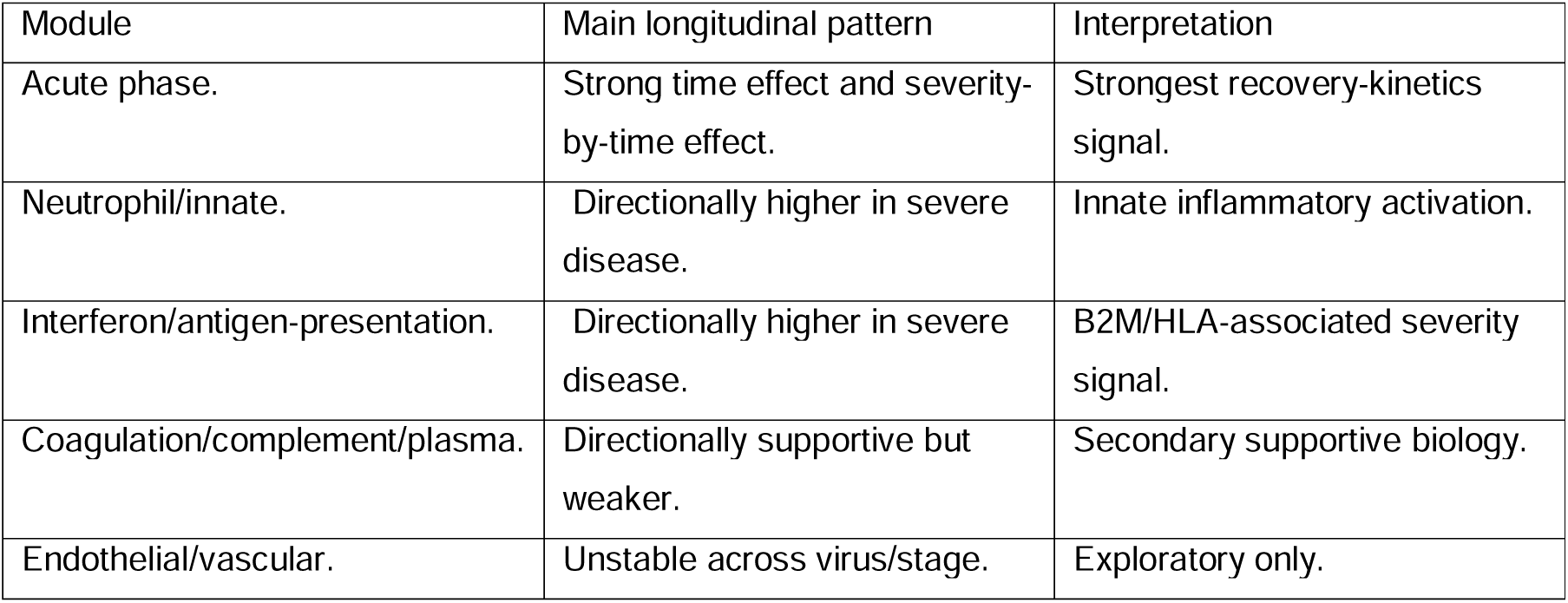
Summary of longitudinal immune-module findings after GEE and recovery-slope analysis.

In conclusion, this study identified a strong and durable plasma proteomic signature of HFRS in humans caused by infection with DOBV or PUUV. The dominant features included acute-phase activation, increased fibrinogen and CRP, later enrichment of immunoglobulin variable-region proteins, and severity-associated perturbation of complement, proteolysis, and platelet-degranulation pathways. These findings support a model in which severe hantavirus disease is shaped by a sustained and dysregulated host response involving inflammation, endothelial dysfunction, complement activation, and platelet/coagulation abnormalities. The candidate severity-associated proteins identified here may ultimately contribute to improved early risk stratification of patients with HFRS.

## Methods

### Ethics Statement

The collection and the use of samples was approved by the Republic of Slovenia National Medical Ethics Committee (69/03/12 and 30/04/15). A written informed consent was obtained by the patient for collection of longitudinal samples. The study was conducted according to the principles expressed in the Declaration of Helsinki.

### HFRS Patients

A total of 50 plasma samples were collected from 13 patients (6 infected with PUUV and 7 infected with DOBV) with confirmed HFRS, hospitalized in different Slovenian hospitals between the years 2010 and 2014. As a control group, 5 healthy adult volunteers (4 males and 1 female) were included. Clinical diagnosis of all patients was laboratory confirmed with serological and molecular tests as described previously ^2, 35, 36^. For each patient a detailed medical chart was collected and based on clinical criteria patients were categorized as severe or mild. Severe infection was defined where a patient had thrombocytopenia <50 x 10^9^/L and the need for dialysis; or thrombocytopenia <50 x 10^9^/L and the presence of at least two of the following: bleeding, oliguria/anuria, and levels of urea and/or creatinine at least 4x higher than the upper normal level. Patients who did not meet these criteria were allocated to the mild disease category. Plasma was also obtained from 6 controls who were seronegative for either DOBV or PUUV (although one failed analysis).

Viral RNA load was measured in patients’ plasma samples using Quantitative One-Step real-time reverse transcriptase polymerase chain reaction (qRT-PCR) as described previously ^37^. Briefly, viral RNA was isolated from patients’ plasma samples using EZ1 Virus Mini Kit v2.0 according to manufacturer’s instructions (Qiagen). For qRT-PCR reaction TaqMan Fast Virus 1-Step Master Mix was used according to manufacturer’s instructions (Applied Biosystems). For DOBV detection primers DOB-S F1 (TTGTTCCTGTTTGCTGGAAAATGAT), DOB-S R1 (CGGGTTGAAGAATGGCTTGAC) and probe DOB-S-MGB-P (FAM-CCGTGCAAGCTAC) were used. For detection of virus PUUV primers PUU D (GGAGTAAGCTCTTCTGC), PUU L (ACATCATTTGAGGACAT) and probe PUU-MGB (VICAGACCAAAGCATTTATATG) were used. qRT-PCR reaction was performed on ABI 7500 Fast instrument (Applied Biosystems) under conditions of 5 min at 50°C, 20 s at 95°C and 45 cycles of 3 s at 95°C, 30 s at 55°C and 30 s at 60°C. DOBV and PUUV RNA was quantified using calibrated synthetic standard gBlock (IDT). A linear effects model was used to track viral load on a per patient basis (log_10_(viral load+1)∼day×virus+day×severity+virus×severity+(1∣patient)

### Proteomic analysis

Plasma samples were heat-treated for 30 minutes at 56 °C before any further processing. Plasma protein concentration was determined using the Pierce™ Bradford Protein Assay Kit (ThermoFisher Scientific). Samples were normalised to 10μg/μl protein with hplc grade water.

### TCA precipitation

Normalised plasma samples were precipitated by adding 8 volumes of ice-cold acetone containing 15% (w/v) TCA, then incubated at -20°C for 2 hours. Samples were centrifuged at 12,000 g for 10 min (4°C) to pellet proteins. Pellets were washed three times with ice-cold acetone and allowed to air dry. Protein pellets were re-suspended in 1 % (w/v) sodium deoxycholate in 50 mM ammonium bicarbonate.

### Tryptic digestion

Proteins were reduced with 3 mM Dithiothreitol (Sigma) at 60°C for 10 min, cooled, then alkylated with 9 mM Iodoacetamide (Sigma) for 30 min at room temperature, protected from light. Proteomic-grade trypsin (Sigma) was added at a protein: trypsin ratio of 50:1 and incubated at 37°C overnight. Samples were acidified to 1% (v/v) with TFA and incubated at 37°C for 2 hours before centrifugation at 12,000 g for 30 minutes at 4°C. The supernatant was retained for MS analysis.

### Nano LC MS ESI-MS/MS analysis

Peptides (0.5μg) were analysed by online nanoflow LC using the Ultimate 3000 nano system (Dionex) coupled with a Q-Exactive HF mass spectrometer (ThermoFisher Scientific). Samples were loaded onto an analytical column (Easy-Spray PepMap® RSLC 50 cm × 75 μm inner diameter, C18, 2 μm, 100 Å, ThermoFisher Scientific). The column was operated at a constant temperature of 30°C. Chromatography was performed with a buffer system consisting of 0.1 % (v/v) formic acid (buffer A) and 80 % (v/v) acetonitrile with 0.1 % (v/v) formic acid (buffer B). The peptides were separated by a linear gradient of 3.8 – 50 % buffer B over 90 min at a flow rate of 300 nl/min followed by a washing step (5 min at 99% solvent B) and an equilibration step (10 min at 3.8% solvent B). The Q-Exactive HF was operated in data-dependent mode with survey scans acquired at a resolution of 60,000. Up to the top 16 most abundant isotope patterns with charge states +2, +3 and/or +4 from the survey scan were selected with an isolation window of 2.0 Th and fragmented by higher energy collisional dissociation with normalised collision energies of 30. The maximum ion injection times for the survey scan and the MS/MS scans were 100 and 45 ms, respectively, and the ion target value was set to 3E6 for survey scans and 1E5 for the MS/MS scans. Repetitive sequencing of peptides was minimised through dynamic exclusion of the sequenced peptides for 20s.

### Data analysis

MS raw data were analysed using FragPipe v23 (https://github.com/Nesvilab/FragPipe), with MSFragger v.4.4.1 ^38^. The built-in LFQ-MBR workflow was used with default settings. Spectra were searched against the human reference proteome downloaded from UniProt (UP000005640_9606, May 2026; 20,670 entries) and two hantavirus reference proteomes (UP000008481_1337063 and UP000202548_3052477, May 2026; 7 entries combined) with a precursor mass tolerance of 20 ppm and fragment mass tolerance of 20 ppm, Mass Calibration and Parameter Optimization was selected. Identifications were filtered to obtain false discovery rates (FDR) below 1% for both peptide spectrum matches (minimum peptide length of 7) and proteins using a target-decoy strategy. For all searches, carbamidomethylated cysteine was set as a fixed modification, and methionine oxidation and N-terminal protein acetylation were set as variable modifications, allowing up to 3 modifications per peptide. Strict trypsin cleavage was set as the protein digestion rule. Label-free quantification was performed using IonQuant v.1.11.20 ^39^. The differential expression analysis was performed in FragPipe Analyst ^40^ using Limma ^41^ and the resulting p-values were adjusted for multiple comparisons using the Benjamini-Hochberg procedure. Max LFQ intensity was used, and the minimum percentage of non-missing values globally (all samples) was set at 50. Perseus style of imputation was utilised. Significant differentially expressed proteins had an adjusted p-value cutoff of 0.05 and a log2 fold change cutoff of 1. In the longitudinal trajectory and module analyses, “abundance” refers to sample-level MaxLFQ protein intensity values. These are relative label-free protein intensity measurements and should not be interpreted as absolute protein concentrations. Fold-change outputs from FragPipe Analyst were used for differential-abundance summaries, whereas the longitudinal trajectory, module-score and recovery-slope analyses used the sample-level protein intensity matrices.

### Data visualisation

Principal component analysis (PCA) was performed using ClustVis ^42^. Unit variance scaling was applied to rows; singular value decomposition with imputation was used to calculate principal components. For analyses using the filtered non-imputed matrix, missingness was considered within relevant virus, severity and time strata rather than only globally across the full dataset. Missing values were not re-imputed at the module-score stage. Module scores were computed from the available detected proteins within each immunologically defined module, and the number or fraction of detected proteins contributing to each score was retained as detection support. Module-level results were interpreted more cautiously when detection support was weak or uneven across virus, severity or time strata. Hierarchical clustering was also performed using ClustVis. Rows were centred; unit variance scaling was applied to rows. Both rows and columns were clustered using correlation distance and complete linkage. Input was filtered to include only significant (adjusted p-value ≤ 0.05, log2 fold change ≥1) early time-point samples for either severe or mild cases from both PUUV or DOBV infections. Highlighted clusters were explored for functional enrichment using ShinyGO v 0.74 ^43^, GO biological processes p-value cut off was set at 0.01, and pathways were visualised as dot plots. All human proteins were used as background. Volcano plots were created using the FragPipe Analyst package. Significant differentially expressed proteins had an adjusted p-value cut-off of 0.05 and a log2 fold change cut-off of 1

### Statistical and longitudinal analysis

Early severity-associated protein differences were assessed using subject-level summaries. Early samples were defined as samples collected within the first four days of hospitalisation. For each protein, repeated early measurements within each subject were collapsed using the median abundance. A severe-minus-mild effect size was then computed as the median of the severe subject-level medians minus the median of the mild subject-level medians. Positive values therefore indicated higher early abundance in severe cases, whereas negative values indicated lower early abundance in severe cases. Exact subject-label permutation tests were used to rank early severity-associated proteins. Because of the small number of subjects and the resulting discrete permutation p-values, these analyses were interpreted as candidate-generating rather than definitive inference. For longitudinal analysis, candidate proteins were grouped into immunologically meaningful modules such as acute-phase inflammation, neutrophil/innate immunity, coagulation/complement/plasma biology, endothelial/vascular biology, and interferon/antigen-presentation biology. For each module, detected protein intensity values were standardised across infected samples and module scores were computed as the median of the available standardised protein values within each sample. Missing values were not re-imputed at this stage. Module scores were interpreted together with detection support, especially for small modules where a score could be driven by a single detected protein.

Exploratory longitudinal models were fitted to assess whether protein or module trajectories differed by disease severity over irregularly sampled time. For each protein or module outcome *y_ij_*, where *i* indexes patient and *J* indexes sample/time point, the main mixed-effects model was:

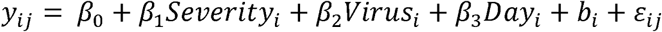

Where bi represents a patient-specific random intercept. An exploratory recovery-kinetics model additionally included a severity-by-day interaction:

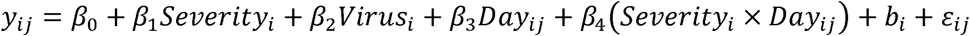

Gaussian GEE models used the same fixed-effect structures but replaced the patient-specific random intercept with patient-level clustering. Given the small sample size, unbalanced sampling across strata, irregular follow-up, and absence of adjustment for clinical time-course covariates, p-values from both mixed-effects and GEE models were interpreted as supportive evidence of effect direction and magnitude rather than formal significance tests.

Patient-level recovery slopes were computed for focal proteins and modules by fitting a simple within-subject linear trend over observed follow-up. These slopes were used as descriptive summaries of recovery kinetics and were not treated as formal group-comparison tests, because they are sensitive to irregular follow-up and to the availability of late severe samples.

## Author contributions

Conceptualization: JAH, TAZ and MK.

Data curation: SA.

Formal analysis: SA, KRR, NK, ED, CEE, IB, TAZ, JAH and MK.

Funding acquisition: TAZ, JAH and MK.

Investigation: SA, KRR, NK and ED.

Methodology: SA, IB, KRR, NK and ED.

Project administration: JAH and MK.

Resources: KRR, NK, TAZ and MK.

Software: SA, CEE, IB and ED.

Supervision: JAH, IB and MK.

Validation: SA, KRR, NK and ED.

Visualization: SA, KRR, NK and ED.

Writing – original draft: SA, ED, CEE, IB, JAH and MK.

Writing – review & editing: All authors.

## Data Availability

All data produced in the present study are available upon reasonable request to the authors
All data produced in the present work are contained in the manuscript

